# Prismatic Effect, Interpupillary Distance and their Correlation with Stereopsis in an African Population

**DOI:** 10.1101/2025.07.05.25330934

**Authors:** Godsway Commanda Osei, Abena Agyeiwaa Owusu, Bernice Kyei-Baffour, Felicity Oppong, Derick Amihere, David Ben Kumah, Clement Afari

**Author notes:** These authors contributed equally to this work (Godsway Commanda Osei, Abena Agyeiwaa Owusu, Bernice Kyei-Baffour, Felicity Oppong, Clement Afari). These authors also contributed equally to this work (Derick Amihere, David Ben Kumah).

## Abstract

**Purpose:** This study aims to explore the relationship between interpupillary distance (IPD), prismatic effects and stereopsis in an African population.

**Methodology:** A descriptive cross-sectional study was carried out with 96 students aged 18 to 35. Participants were recruited using a non-probability convenience sampling method. IPD was measured using a pupillometer, while stereopsis was assessed with the Randot stereotest. Data were analysed using IBM SPSS Statistics version 27.0.1, with a significance level set at p ≤ 0.05.

**Results:** The study included 96 participants: 32 males (33.3%) and 64 females (66.7%), with a mean age of 21.61 (±1.57) years. The age range was 18 to 35. The findings showed that a wider near IPD was associated with poorer stereopsis (p < 0.001). Additionally, introducing prisms in front of the eyes led to a significant reduction in stereopsis (p < 0.001). A significant difference in stereopsis was found between males and females (p = 0.02), with males having a mean stereopsis of 30.16 (±7.01) and females 26.72 (±6.80). There was also a significant difference in near IPD between genders (p < 0.001), with males averaging 64.7 (±3.34) and females 61.5 (±2.06).

**Conclusion:** Smaller near IPD is associated with better stereopsis. It also demonstrated that prismatic effects, whether vertical or horizontal, significantly impact local stereopsis, emphasizing the importance of accurate IPD measurement. Females were found to have a smaller IPD and better stereopsis compared to males.

## Introduction

The human visual system is a complex system that plays an essential role in our daily lives [1], facilitating our perception and interaction with the environment. A key feature of the visual system is depth perception, which enables us to ascertain the relative distance of objects in our surroundings [2]. Depth perception is achieved through a process called stereopsis, which involves the integration of visual information from both eyes [2]. Stereopsis is considered the most powerful cue to depth perception [3].

Stereopsis results from horizontal retinal image disparity between the two foveae or other corresponding retinal points [4]. This disparity enhances the human visual system’s ability to perceive depth through binocular vision. [4]. In essence, our depth perception improves when each eye views slightly different images due to their positions, and the brains’s visual cortex processes these differences referred to as horizontal or binocular disparities, to producw the sensation of depth [4–7].

Several factors influence stereoacuity, including age, gender, vergence, viewing distance of the target and monocular cues [8]. While monocular cues can also provide depth perception, they are not as precise or vivid as stereopsis, which offers an immersive and distinct depth experience [7].

Stereoacuity, which measures the smallest disparity that an individual can detect, is a key indicator of the quality of stereopsis [9]. The smallest angle of binocular disparity (measured in arc seconds) that produces the perception of depth is known as threshold stereoacuity [10, 11].

Stereopsis is particularly advantageous in tasks involving complex visual presentations and hand eye coordination[12–14]. It plays a very important role in practical activities such as needle-threading, catching fast-moving balls, pouring liquids, and opetrating stereoscopic instruments like binocular microscope [13]. Additionally, as 3 dimension display technology becomes increasingly prevalent in entertainment, medicine, and scientific imaging, possessing high-quality binocular vision, including stereopsis, is becoming more important in modern society [14].

Stereopsis depends on an intact binocular vision system, which requires both eyes to work together effectively [12]. Interpupillary distance, the distance between the centers of the pupils in each eye is an important anatomical parameter in binocular vision [8]. IPD varies by gender age, and ethnicity, and it differs between near and distance viewing conditions [15]. These variations may influence the quality of binocular vision and, by extension stereopsis [15, 16]. Among the many factors that affect stereopsis, IPD is the only one that can be actively manipulated to enhance stereoacuity [8, 17]. This highlights the significance of IPD in optimizing stereoscopic function. IPD determines the degree of retinal image disparity, which the brain combine to generate depth perception [8, 18].

In the optical industry, precise IPD measurement is very important for proper eyeglass fitting. An incorrect IPD measurement during the fitting of ophthalmic lenses can lead to decentration, which induces unintended prismatic effect [8, 15]. A Prismatic effect refers to the image displacement that occurs when light passes through a prism [15]. Similarly, when light passes through an ophthalmic lens other than the optical center, the lens can act like a prism and displace the image [15, 19]. These unintended induced prisms can alter the alignment of the visual axes and potentially affect depth perception [19, 20].

This study, therefore, aims to explore the intricate relationship between interpupillary distance and stereopsis, and to examine the effects of unintended induced prismatic deviations on stereopsis. By establishing these connections, eye care professionals can gain a better understanding of how induced prisms influence stereopsis and can incorporate IPD considerations when prescribing lenses.

In many developing countries, spectacles are widely used to address a range of visual problems from refractive errors to eye protection, largely due to their affordability and ease of use. However, a common and easily made error during spectacle fitting is the induction of unintended prisms, which may negatively affect stereopsis.

Limited research has been conducted on the correlation between interpupillary distance and stereopsis in African populations. Factors such as race have been identified as influencing our IPD, suggesting a possible link between racial demographics and the characteristics of stereopsis [8]. Racial disparities in physiological traits such as facial structures and ocular anatomy, indicates that variations in IPD across racial groups may influence depth perception [8, 21]. Therefore, while previous studies have primarily focused on non-African populations, there is a pressing need to include individuals of African descent and other underrepresented groups in such research.

By examining these factors in tandem, this study seeks to elucidate their individual and collective contributions to binocular vision, thereby offering insights into how spectacle prescription practices can be optimized to improve patient. Overall, this research endeavours to address existing methodological gaps by conducting a comprehensive investigation into interpupillary distance, unintended prismatic effects, and their impact on stereopsis within a specific setting. By addressing these gaps, we aim to contribute valuable insights that can inform clinical practices and improve patient care outcomes.

### Literature review

IPD’s clinical significance lies in aiding the accurate placement of eyeglass lenses in front of the eyes, preventing strain caused by prismatic effects from the lenses. To correctly fit ophthalmic lenses, specific points on the lenses must align with the center of each eye’s pupil for far and near viewing. IPD measurement methods include manual measurement with a PD

Rule or digital measurement with a pupilometer. Using a pupilometer offers the advantage of more precise measurement of monocular PDs, which is especially useful for ordering lenses for high refractive errors or progressive addition lenses requiring accurate centration [15].

A cross-sectional study conducted by Kumah et al in 2016 to measure the interpupillary distance (IPD) of students in Kumasi Metropolis involving 500 students aged 10 to 20 from Junior and Senior High Schools in Kumasi Metropolis. IPD was measured using both a pupilometer and a PD rule. Among the 500 students, 290 (58%) were males. The study found that using a pupillometer, the average distance and near IPD for students were 65.53 ± 3.348 mm and 61.60 ± 3.054 mm, respectively [15]. When measured with a PD rule, the IPD was 64.48 ± 3.429 mm for distance and 62.01 ± 3.464 mm for near vision. It was further stated that Similarly, a study by Eom et al in 2013 reported a significant difference in IPD between male and female subjects (p=0.014), with males having an IPD that is, on average, 2 mm larger than females, with a standard deviation of ±1.5 mm [22]. This study also reports that interpupillary distance (IPD) is the only component of stereopsis which can be manipulated to enhance our stereopsis [22].

A study by Zaroff et al in 2003, reported a significant positive correlation between age and stereoacuity (r = +0.44, p < 0.0001) [23]. Zaroff and colleagues also highlighted that age had a statistically significant influence on stereoacuity thresholds (F = 13.58, p < 0.0001). They also noted that one key factor contributing to decreased stereoacuity in older adults is senile miosis, a condition where pupillary size decreases with age, leading to a 44% reduction in retinal illuminance and, consequently, a decline in retinal sensitivity to visual stimuli [23].

Similarly, Garnham et al in 2006, explored the effect of age on stereoacuity, using four different stereotests to measure stereopsis in 60 participants aged 17 to 83. Their findings showed a significant decline in stereoacuity with age across all stereotests (p < 0.001 for all) [24]. They also observed a reduction in stereoacuity between the age groups of 30-49, 50-69, and 70-83, as measured by TNO and Frisby near stereotests (r = 0.55 and r = 0.62, respectively) [24].

Laframboise et al in 2006, also reported a significant but moderate correlation between age and stereoacuity (r = 0.33; p < 0.01), explaining that normal aging affects binocular correlation processing [25]. The correlation between age and stereopsis was low but significant (r = 0.33; t98 = 3.27; p<0.01) and yielded a determination factor of 11%. [25].

A study in 2014 by Jafari et tal, reported a statistically significant difference in stereoacuity between males and females (p= 0.015). Concluding that females have better stereoacuity compared to male subjects [8].

Some studies have explored the relationship between IPD and stereoacuity with varying methodologies and samples.

A study by Jafari et al, examined the relationship between IPD and stereoacuity. The results showed that individuals with smaller IPD have better stereoacuity threshold and that female subjects have both smaller IPD and better stereoacuity compared to those with wider IPD which is mostly common in male subjects [8]. However, a study conducted in 2013 by Eom et al., investigated the effect of IPD on stereoacuity in a large sample of participants. They found a positive correlation between IPD and stereoacuity, suggesting that individuals with a larger IPD tend to have better stereoacuity [22]. The conflicting findings in the literature may be attributed to various factors such as age, sample size, and measurement techniques. Additionally, differences in measurement techniques, such as the use of different stereoacuity tests, may contribute to the discrepancies observed in the literature.

Most literatures indicate that individuals with smaller IPD tend to have better depth perception and 3D vision. However, conflicting findings and methodological limitations highlight the need for further research in this area.

In another study by Kester et al, investigating interpupillary distance and Stereoscopic threshold, eight optometry students participated voluntarily, undergoing changes in IPD via a head-mounted device. The subjects’ natural IPD and artificial IPD were measured using an optical apparatus. Stereo-threshold was assessed using a Howard-Dolman apparatus positioned 3 meters from the subject. Subjects adjusted two pegs until they perceived them as parallel, with twenty settings made for each IPD condition. Analysis revealed significant differences in Stereo-threshold across the three IPD conditions. Their findings suggested that Stereo-threshold is not constant when IPD is artificially increased. It was therefore concluded that contrary to the notion that Stereo-threshold remains consistent irrespective of IPD, the results indicate variability in stereoscopic perception with IPD alterations [26]. This underscores the importance of considering individual differences in IPD when assessing stereovision.

A study by Rènée du Toit et al., on tolerance to prism induced by readymade spectacles reported that highest prism powers (1 prism BU, 2 prism BO, 2 prism BI) could not be worn for 8 hours by the majority of participants [27]. Comfort ratings at near (similar to those at distance) were statistically significantly different when the highest prism power was compared with each of the lower powers (vertical prism: both the control and 0.5 Delta differed from 1 Delta; horizontal prism: the control, 0.5 Delta and 1 Delta all differed from 2 Delta). This poses a need to assess the direct effect of such high prism powers on stereopsis [27].

A study by Momeni-Moghaddam et al., to assess the effects of vertical prism diopters on local and global stereopsis in a sample of 99 participants. Their findings revealed a significant reduction in both local and global stereopsis thresholds when vertical disparity was induced, particularly in cases where the prism was placed in front of the non-dominant eye [28]. They examined variations in stereopsis thresholds under different conditions of induced vertical disparity. Their results indicated a notable discrepancy between local and global stereopsis thresholds, with global stereopsis being more adversely affected by induced vertical disparity [28].

A study by Arshad et al., investigated prevalence of decentred spectacles among population group and explored their impact on various visual parameters. Horizontal and vertical decentration were assessed, revealing a high prevalence of misalignment in both directions [19]. The findings indicated that a significant proportion of individuals exhibited horizontal and vertical decentration, with notable discrepancies between interpupillary distance (IPD) and optical centration distance (OCD). The study identified a predominance of base-in prisms induced in spectacles, particularly in the vertical direction. Horizontal prismatic effects were correlated with a decline in stereoacuity, with a substantial number of individuals experiencing reduced stereoacuity within specific ranges of prismatic effects. The study concluded that improper dispensing of spectacles contributes to the induction of prismatic effects, leading to a shift in image position on the retina and subsequent reduction in stereoacuity. These findings underscore the importance of accurate spectacle dispensing and highlight the need for further research to mitigate the impact of decentred spectacles on visual function.

## Materials and methods

### Study design

A descriptive cross-sectional study was employed. Data were collected on participants’ interpupillary distance (IPD) and baseline stereopsis. Prism was then induced, and the resulting change in stereopsis was remeasured at a single point in time. This provided useful insights into the nature of the population, as well as the relationship between IPD, prismatic effect and stereopsis. The study was conducted in Kumasi, the capital city of the Ashanti region of Ghana, specifically at the Kwame Nkrumah University of Science and Technology (KNUST). The Optometry clinic at the College of Science served as the primary site for data collection. Convenience sampling was used to recruit participants to meet the required sample size. Participants were selected based on their availability and willingness to participate in the study.

### Data collection tools

A LOGMAR visual acuity chart, autorefractor, Randot Stereoacuity booklet, Polarised glasses, phoropter, Welch Allyn direct ophthalmoscope were used in the study. Additionally, a standardized questionnaire was employed to assess the fatigue levels of the individual participants. The questionnaire consisted of closed-ended questions and was thoroughly explained to participants who did not fully understand it. Patients’ demographics as well as their past medical and ocular history, were also collected. Demographic data included age, sex, programme of study and other relevant details. Information related to the use of glasses, date of last eye exam, drug use, other health conditions, amblyopia and strabismus was also obtained.

To ensure validity and reliability of data collection tools, the following were implemented: A standardized questionnaire was used, a pilot study was conducted at the beginning of the research, researchers who comprised of optometrists, intern optometrists and final year optometry students were trained prior to data collection to minimize measurement errors. Specific roles were assigned to each researcher to reduce inter observer variability. The test-retest method was employed to obtain consistent results. A calibrated pupillometer was used to accurately measure participants’ interpupillary distance (IPD) at both near and distance. Induced prismatic effects were carefully controlled using consistent amounts of base-in, base-out, base-up, and base-down prisms. Stereopsis was measured both before and after prism induction. The assessment of stereopsis was conducted using a reliable clinical tool (Randot Stereotest), ensuring consistency across all measurements.

### Data collection

The study was conducted in accordance with the Declaration of Helsinki and received ethical clearance from the Committee on Human Research, Publication, and Ethics at the School of Medical Sciences, KNUST (CHRPE/AP/276/24). Data was collected at the Optometry Clinic, at the College of Science building, KNUST from May to July 2024. Written informed consent was obtained from all participants, and each procedure was thoroughly explained to them during the examinations. Participants’ rights, privacy, and confidentiality were ensured. Participants also had the right to withdraw from the study at any time without any consequences.

Inclusion criteria included students aged 18 to 35 years with best-corrected visual acuity (BCVA) of 0.2 LogMAR or better. Exclusion criteria included students with visual acuity worse than 0.2 LogMAR, strabismus, phoria greater than ±3 prism diopters, stereoacuity worse than 60 seconds of arc, any manifest deviation (congenital or post-surgical), blind eyes or any external or internal eye disease.

Visual acuity was assessed at both distance (6 meters) and near (40 centimeters) using the LogMAR visual acuity (VA) chart. Proper illumination was used, and participants were given appropriate instructions to ensure accurate measurements. Participants who wore spectacles were tested with and without their spectacles to read with their spectacles, and results recorded accordingly. Visual acuity was assessed monocularly and then binocularly. The participants were instructed to read the optotypes on the VA chart, and the smallest line read correctly was recorded in LogMAR notation.

A cover test was performed to detect any manifest or latent ocular deviation at near using a handheld occluder and a measuring prism. Both the cover-uncover test and the alternating cover test were conducted. The cover-uncover test was done to rule out tropia, and the alternating cover test was used to detect phoria.

The anterior and posterior segment of the eye were examined using the ophthalmoscope. Participants were instructed not to look directly at the ophthalmoscope light during the assessment and were reassured of the safety of the procedure. Any clinical findings were recorded on the data collection sheet.

Objective refraction was performed for all participants using an autorefractor. Each participant was seated behind the autorefractor with their forehead and chin properly positioned on the rest. A researcher operated the device to obtain the measurements. The results from the objective refraction were then refined using a phoropter to determine the participants’ best corrected visual acuity. Dry refraction was conducted, and accommodation was controlled using a high plus lens during subjective refraction.

Participants who did not meet the predetermined criteria based on visual acuity, cover test results, ophthalmoscopy findings and objective refraction outcomes were excluded from the study. Participants who met the inclusion criteria were given a validated fatigue assessment questionnaire to evaluate their fatigue levels.

Interpupillary distance was measured using a digital pupillometer (model: PD-82II). Measurements were recorded in millimeters. Each participant was seated at eye level with the examiner. The pupillometer was gently placed over subject’s eyes with the forehead support in place. The nose pads were properly positioned, and the forehead bar helped to center the instrument.

The examiner looked through the eyepiece while the participant focused on the fixation target. The examiner aligned the crosshairs with the participant’s corneal reflex, and the IPD was recorded as the average of three readings in millimeters(mm);1. The distance wheel was set to infinity (∞) for far IPD and to 40 cm for near IPD; 2. Both monocular and binocular IPDs were measured and recorded; 3. Measurements were repeated three times to ensure accuracy and consistency. The final recorded value represented the distance between the centers of the pupils.

Stereopsis was measured using the Randot stereotest. Participants wore polarized glasses, and ametropes used their spectacle correction with the polarized glasses positioned in the phoropter during testing. Each participant was asked to identify six geometric shapes from a distance 40 cm. Disparity was measured in seconds of arc, with testing targets ranging from 800-40 seconds of arc. Additional lighting was provided to ensure consistent illumination for all participants during the test.

Horizontal and vertical prisms were used to induce prismatic effect in each participant. Prisms were introduced in front of the participant’s eyes, and stereoacuity thresholds were remeasured. Specifically, 2 prism diopters were induced in base-in and base-out directions, while 1 prism dioptre was used for base-up and base-down directions. After prismatic induction, stereoacuity was measured. Horizontal prisms were introduced first for all participants. A suppression check was performed after prism induction to identify and exclude any participants suppressing input from either eye.

## Statistical analysis

All the data was entered in excel and cleaned. Afterwards they were coded and exported into IBM SPSS Statistics version 27.0.1

The study utilized IBM SPSS Statistics version 27.0.1 to conduct an in-depth analysis of the study population and find the relationship between various parameters. Inferential and descriptive statistics, encompassing measures such as mean, standard deviation and frequency distribution, were employed to provide a comprehensive overview of the data. Shapiro Wilk’s test was used to check whether the data was normally distributed or not. Furthermore, the relationship between interpupillary distance and stereopsis was investigated using Spearman’s rho which allowed for a nuanced understanding of their interrelationships. Comparison of baseline and induced stereopsis were analyzed with Wilcoxon Signed Ranks Test, comparison of sex against stereopsis and interpupillary distance was analyzed with Mann-Whitney U test, however, comparison of age and stereopsis was analyzed with Kruskal - Wallis test.

## Results

### Socio-demographic

A total of 96 students passed the criteria set and were involved in the study. Of these, 32 (33.3%) were males and 64 (66.7%) females. The mean (±SD) age was 21.61 (±1.57) years. Majority of the students studied Doctor of Optometry, 53 (55.2%) and were in their fourth year, 53 (55.2%). All the participants were students in the College of Science. Table 1 provides more information on the students’ demographics.

**Table 1.**
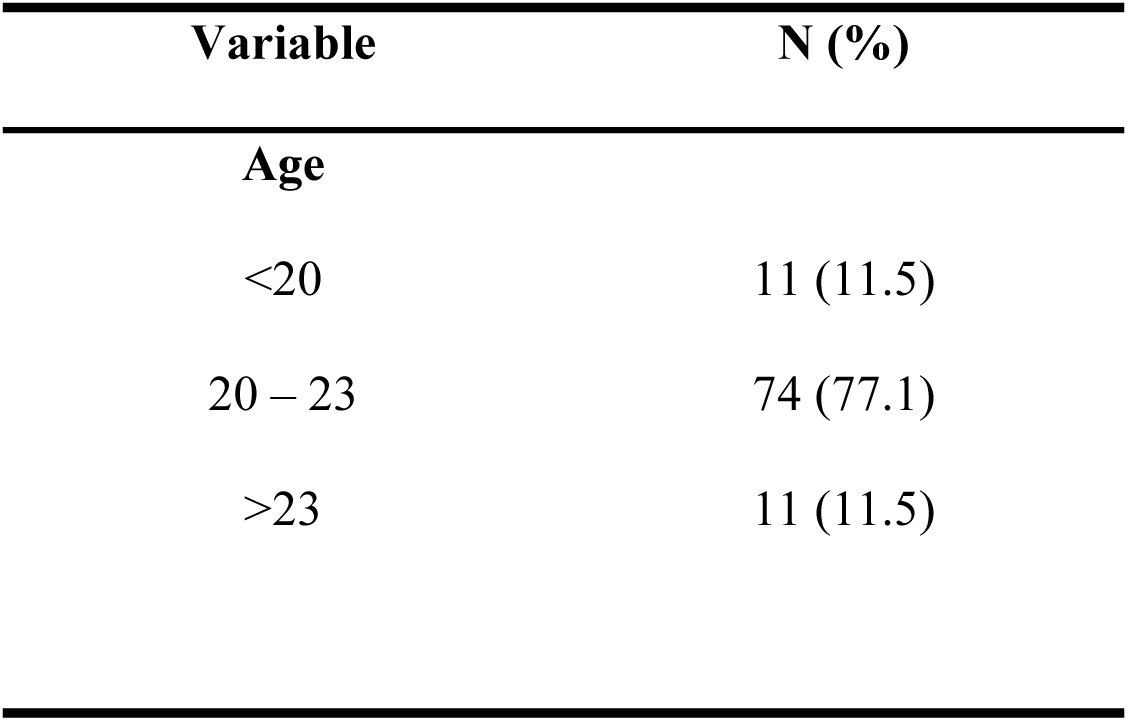
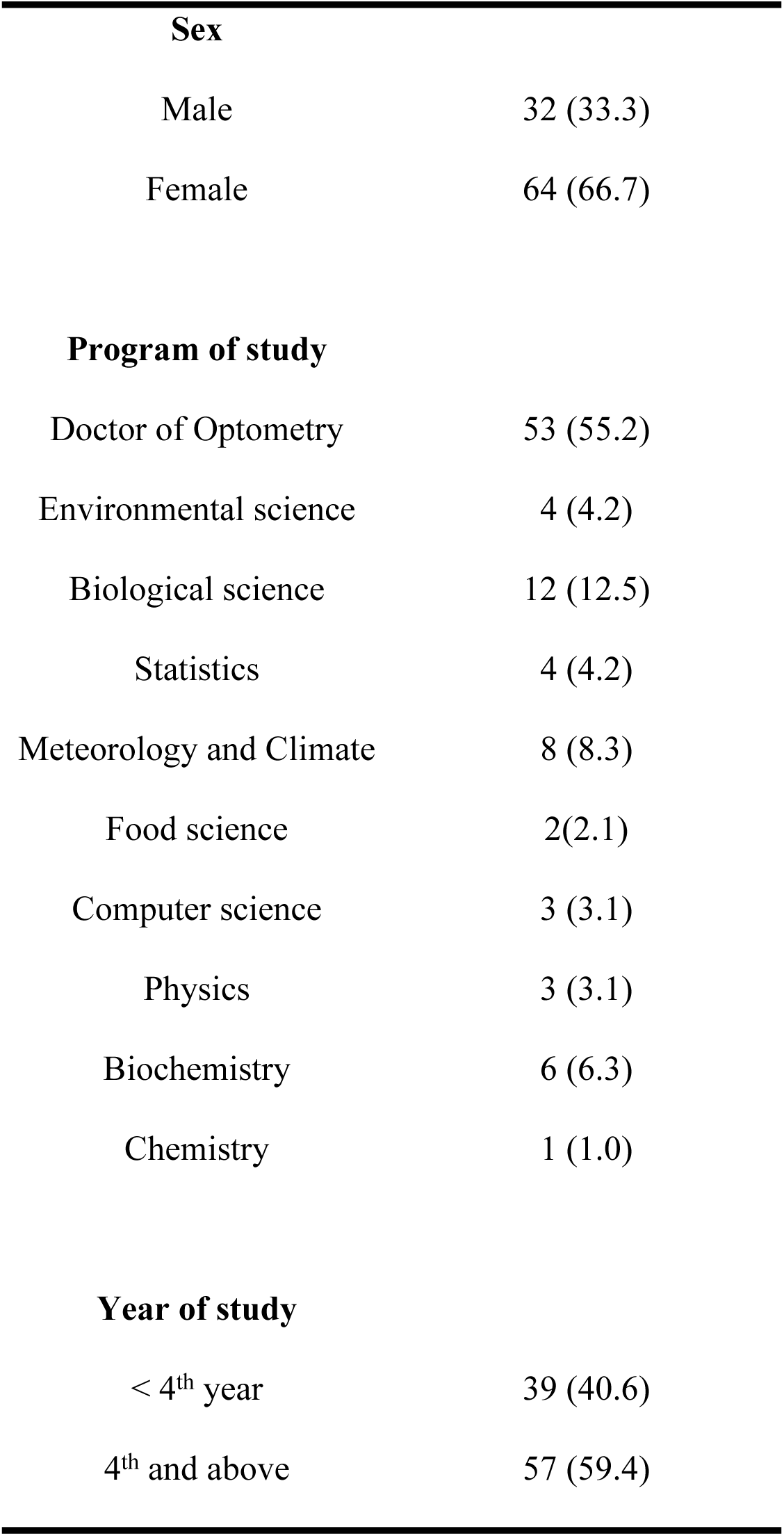
Students Demographics.

| Variable | N (%) |
| --- | --- |
| **Age** |  |
| <20 | 11 (11.5) |
| 20 – 23 | 74 (77.1) |
| >23 | 11 (11.5) |

|  |  |  |
| --- | --- | --- |
| 338 | **Sex** |  |
| 339 | Male | 32 (33.3) |
| 340 | Female | 64 (66.7) |
| 341 |  |  |
| 342 | **Program of study** |  |
| 343 | Doctor of Optometry | 53 (55.2) |
| 344 | Environmental science | 4 (4.2) |
| 345 | Biological science | 12 (12.5) |
| 346 | Statistics | 4 (4.2) |
| 347 | Meteorology and Climate | 8 (8.3) |
| 348 | Food science | 2(2.1) |
| 349 | Computer science | 3 (3.1) |
| 350 | Physics | 3 (3.1) |
| 351 | Biochemistry | 6 (6.3) |
| 352 | Chemistry | 1 (1.0) |
| 353 |  |  |
| 354 | **Year of study** |  |
| 355 | < 4<sup>th</sup> year | 39 (40.6) |
| 356 | 4<sup>th</sup> and above | 57 (59.4) |
| 357 |  |  |

### Relationship between Near IPD and stereopsis

The relationship between near IPD and stereopsis (local and global) was analyzed using non parametric correlation. There was statistically significant difference in local stereopsis between the different near interpupillary distance, p<0.001 (Table 2). However, there was no statistical significance in global stereopsis and distance IPD, p>0.05. This is illustrated in table 2.

**Table 2:** Relationship between near IPD, local stereopsis.

| Spearman's rho |  | NIPD | Local stereopsis |
| --- | --- | --- | --- |
| NIPD | Correlation Coefficient | 1.000 | .670** |
|  | Sig. (2-tailed) | . | <.001 |
| Local stereopsis | Correlation Coefficient | .670** | 1.000 |
|  | Sig. (2-tailed) | <.001 | . |

The results indicate that as near IPD increases, the value of local stereopsis also increases which means, stereopsis worsens as near IPD increases. The scatter plot (Fig 1) provides a regression formula for calculating local stereopsis score based on near IPD. Y=76.33+1.67X;

**Fig 1:**
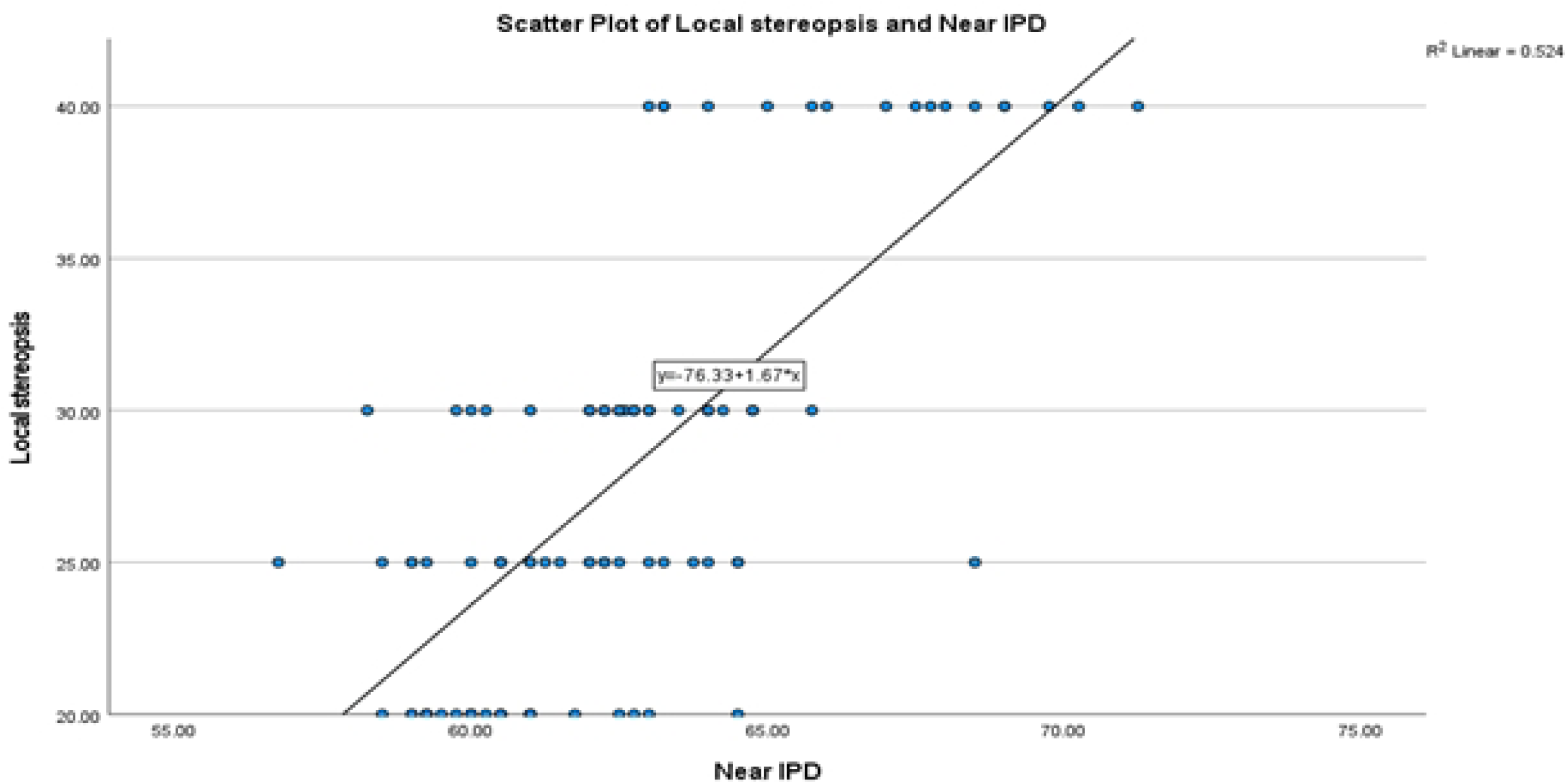
Relationship between near interpupillary distance and local stereopsis. Where Y represents the stereopsis values and X represents the near IP

### Relationship between near IPD and global stereopsis

The results indicate that there is no statistically significant difference between near IPD and global stereopsis. More information is provided in table 3.

**Table 3:**
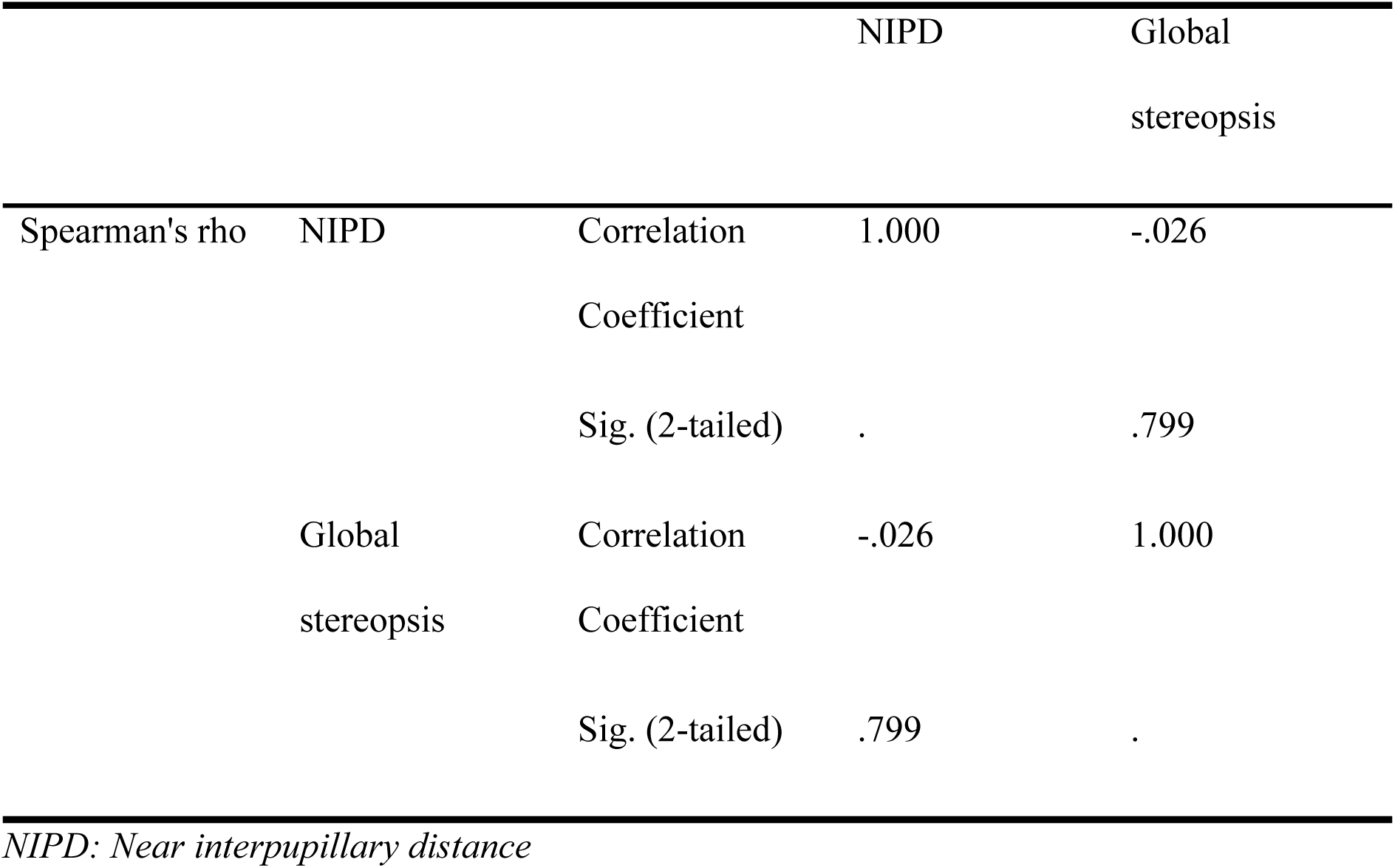
relationship between near interpupillary distance and global stereopsis.

### Comparison of baseline and prism induced stereopsis

Wilcoxon Signes-Rank Test was conducted to compare the medians of the baseline local stereopsis and induced local stereopsis (base in, base out, base up and base down). The test showed a statistically significant difference between the medians of baseline stereopsis and base out, base in, base up and base down induced stereopsis. Table 3 provides more details about it.

**Table 3:** comparison of baseline and prism induced stereopsis.

| Groups | Median | Paired groups | Z score | P - value |
| --- | --- | --- | --- | --- |
| A | 25.00 |  |  |  |
| B | 40.00 | A-B | -8.264 | <0.001* |
| C | 50.00 | A-C | -8.167 | <0.001* |
| D | 40.00 | A-D | -8.211 | <0.001* |
| E | 50.00 | A-E | -8.372 | <0.001* |

### Comparison of sex and local stereopsis

This was assessed using non parametric correlation, Mann-Whitney U test, which gave a statistically significant value (p=0.02). The mean (SD) local stereopsis for males and females were 30.16 (SD=7.01) and 26.72 (6.80) respectively. Figure 2 provides details of the model.

**Fig 2:**
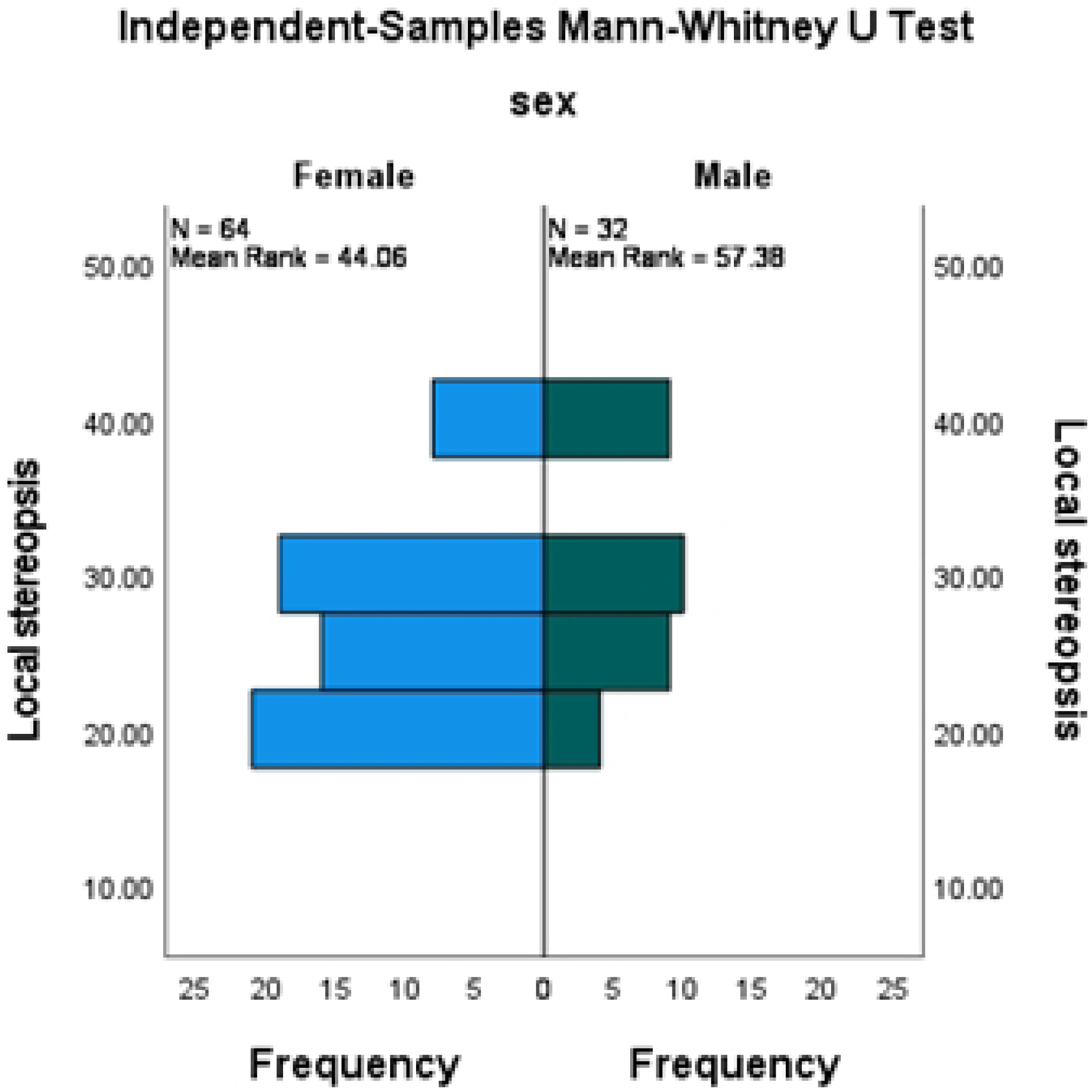
stereoacuity analysis based on sex

### Comparison of sex and near IPD

Mann-Whitney U test revealed there was a statistically significant relationship between sex and near IPD (p<0.001). Table 4 provides information on the mean and standard deviations of near IPD.

**Table 4:** The mean and standard deviations of near IPD for both sexes.

| Sex | N | Mean | Std. Deviation | p-value |
| --- | --- | --- | --- | --- |
| Male | 32 | 64.7 | 3.34 |  |
| Female | 64 | 61.5 | 2.06 | <0.001 |

### Comparison of age and local stereopsis

Results using non parametric correlation, Kruskal-Wallis test showed there was no difference in local stereopsis across the various age groups (P=0.06).

**Table 4:** Stereoacuity analysis based on age.

|  | N | Mean | Std. Deviation | p-value |
| --- | --- | --- | --- | --- |
| <20 | 11 | 32.7273 | 7.53778 |  |
| 20-23 | 74 | 27.0270 | 6.45951 | 0.06 |
| >23 | 11 | 28.6364 | 6.74200 |  |

### Comparison of fatigue and local stereopsis

The results using non parametric correlation, Kruskal-Whitney test, showed that fatigue had no statistically significant impact on stereopsis (P=0.67).

**Table 5:** Stereoacuity analysis based on fatigue level.

| Fatigue level | N | Mean | Std. Deviation | p-value |
| --- | --- | --- | --- | --- |
| severe | 2 | 30.0000 | .00000 |  |
| mild - moderate | 36 | 27.6389 | 6.81240 | 0.67 |
| normal | 58 | 27.9310 | 6.94808 |  |

## Discussion

A total of 96 students within the age range of 18 to 35 years were involved in the study. Of the participants, 32 (33.3%) were male and 64 (66.7%) females. The mean age was 21.61 years (± 1.57 SD). The majority of the students were pursuing Doctor of Optometry degree, 53 (55.2%) participants and were in their fourth year of study.

To determine the relationship between interpupillary distance and stereopsis, the study found a statistically significant correlation between near IPD and stereoacuity (p = 0.001). Additionally, it was established that subjects with smaller IPD had better stereoacuity threshold. This finding is consistent with the results of Jafari et tal, who also reported a statistically significant relationship between near IPD and stereopsis (p = 0.013). A remarkable explanation is that a shorter near IPD reduces binocular disparity, making it easier for the brain to fuse images, thereby resulting in finer stereopsis [29].

However, the findings in this study partially contradicts those of Eom et al. These discrepancies highlight the nuanced effects that IPD may have on stereoacuity under different testing conditions. In this study, both distance and near IPD were measured, and fine stereopsis was assessed at a fixed near distance of 40 cm using the Randot stereotest. In contrast, Eom et al., evaluated the effect of IPD on stereoacuity using two distinct tests: the Frisby Davis Distance (FD2) stereotest, which stimulates real-world distance viewing, and a 3D monitor-based distance stereotest. The FD2 demonstrated that increasing the IPD led to improved stereoacuity. Participants’ stereoacuity thresholds improved when their IPD was artificially increased two-and threefold, suggesting that in real-world environments, a larger IPD enhances depth perception by increasing binocular disparity.

In contrast, the 3D monitor-based stereotest, a simulated stereoscopic environment, yielded opposite results. In this setting, as IPD increased, stereoacuity worsened. The ability to detect fine stereoscopic details decreased with a larger artificial IPD, indicating that in stimulated environments, increased IPD may impair depth perception.

The findings in this study align closely with the results of Eom et al.’s 3D monitor-based stereotest. Using the Frisby near stereoacuity test, it was found that participants with smaller IPD detected finer disparities than those with larger IPD, further contrasting with the FD2 outcomes.

To compare stereopsis before and after prism induction, this study found a significant reduction in local stereopsis when both horizontal and vertical prisms were introduced (p < 0.001). This result is consistent with findings by Momeni-Moghaddam et al., who reported a significant reduction in both local and global stereopsis (p < 0.001) following the induction of 1 prism dioptre Δ of vertical prism. They observed that local stereopsis thresholds worsened by an average of 10 seconds of arc or less under these conditions.

In contrast, this study did not observe a statistically significant reduction in global stereopsis after induction of vertical and horizontal prisms (p > 0.05). Contradicting Momeni-Moghaddam et al.’s report, which indicated a reduction of over 100 seconds of arc in global stereopsis with the same 1.0 Δ of induced vertical prism. This discrepancy may stem from the limited sensitivity of the global stereopsis test this study used, which only measure up to 250 seconds of arc.

The findings of this study are also in agreement with Arshad et al., who reported that prismatic effects, caused by improper fitting of glasses that shift the retinal image, led to reduced stereoacuity. They noted that horizontal prismatic effects in the right eye resulted in a decline in stereoacuity (p = 0.019). The largest group of participants (42) experienced a reduction in stereoacuity within the range of 20-100 seconds of arc when exposed to horizontal prismatic effect between 0 to 1.49 Δ in the right eye.

This study also examined stereopsis in relation to demographic variables and observed two key findings: (1) a significant difference in IPD between sexes and (2) a significant relationship between sex and stereoacuity. Specifically, female participants had a significantly smaller near IPD compared to males (p=0.001), with means of of 61.5mm (±2.06) for females and 64.7mm (±3.34) for males. This is consistent previous research by Kumah et al., (2016) who reported near IPD means of 62.07 ± 3.082 mm for males and 60.95 ± 2.899 mm for females, attributing to the difference to larger craniofacial skeleton in males. Jafari et al., also reported mean near IPD of 61 ± 6.5 mm for males 57 ± 4 mm for females.

This study found a statistically significant difference in local stereoacuity between sexes (p = 0.02), with females demonstrating better stereoacuity than male sub. This finding is consistent with a study by Jafari et al (2014), who also reported a statistically significant difference in stereoacuity between genders (p = 0.015).

In evaluating stereoacuity across three age groups (18-20, 20-23, 23-29,) this study found no significant differences among the groups (p=0.06). This contrasts with findings by Zaroff et al (2002), who reported a significant positive corelation between age and stereoacuity (r = +0.44, P < 0.0001), along with significant effects of age on stereoacuity thresholds (F = 13.58, p < 0.0001). This study’s selected age range of 18-35 years was chosen to reflect the optimal period for stereopsis and to monitor changes following prism induction. The lack of significant age-related findings in this study may be attributed to the narrower age range and exclusion of older individuals. Zaroff et al. may be explained by our narrower age range and the exclusion of older participants. Zaroff et al. suggested that age related declines in stereoacuity could be due to senile miosis, which reduces pupil size, thereby decreasing retinal illuminance and sensitivity by 44%.

Similarly, Garnham et al., examined the effect of aging on stereoacuity in 60 normal participants aged 17-83 years. They found significant stereoacuity decline across all four stereotests used (p < 0.001), particularly between the 30-49 and 50-69 age groups, and again between the 50-69 and 70-83 age groups. These findings point to a marked decline in stereoacuity after age 40. Laframboise et al., also reported that normal aging leads to a statistically significant decline in binocular correlation processing, which was weakly but significantly associated with stereoacuity (r = 0.33; t(98) = 3.27; p < 0.01), accounting for 11% of the variance.

Finally, this study examined the relationship between fatigue levels and habitual stereoacuity. No statistically significant correlation was found (p = 0.67), likely most participants experienced only normal to mild or moderate levels of fatigue.

### Recommendations

Future studies should include a wider age range of participants, particularly older adults to better understand the impact of age on stereoacuity and to assess age-related changes. This approach would help determine how variations in IPD associated with aging influence stereoacuity over time. Expanding the participant pool would allow for a more comprehensive understanding of how aging affects stereopsis and could validate the findings of previous research across a broader population. Additionally, incorporating a variety of stereoacuity tests and viewing distances may yield deeper insights into the factors that influence depth perception. Longitudinal studies would also be beneficial, as they could track changes in stereoacuity and IPD over time, offering valuable information on developmental trends and potential interventions.

## Conclusion

This study reveals the significant relationship between interpupillary distance, prismatic effects, and stereopsis, with gender-based variations observed in both interpupillary distance and stereopsis. The findings indicate that individuals with smaller IPD tend to have better depth perception, while prismatic effects negatively impact stereopsis. Understanding these relationships contributes to improved clinical management of binocular vision issues, particularly in relation to prism use. The study also recommends future research involving a broader age range, varied testing conditions, and longitudinal designs to further investigate the effects of aging on interpupillary distance and stereopsis. Such research would provide a more comprehensive framework for the assessment and management of visual perception.

## Disclosure

This research did not receive any grant from funding agencies in the public, commercial, or not-for-profit organizations.

## Declaration of Competing Interest

The authors have no conflicts of interest to declare.

## Data Availability

All relevant data are within the manuscript and its supporting information files

## Notes

### Competing Interest Statement

The authors have declared no competing interest.

### Funding Statement

The author(s) received no specific funding for this work.

### Author Declarations

The study was conducted in accordance with the Declaration of Helsinki and received ethical clearance from the Committee on Human Research, Publication, and Ethics at the School of Medical Sciences, Kwame Nkrumah University of Science and Technology. Written informed consent was obtained from all participants, and each procedure was thoroughly explained to them during the examinations. Participants‘rights, privacy, and confidentiality were ensured. Participants also had the right to withdraw from the study at any time without any consequences.

